# Prevalence and risk factors associated with malaria infections at a micro-geographic level in three villages of Muheza district, north-eastern Tanzania

**DOI:** 10.1101/2024.03.04.24303697

**Authors:** Daniel P. Challe, Filbert Francis, Misago D. Seth, Juma B. Tupa, Rashid A. Madebe, Celine I. Mandara, Emmanuel S. Kigadye, Vedastus W. Makene, Deus S. Ishengoma

**Affiliations:** National Institute for Medical Research, Tanga, Tanzania; The Open University of Tanzania, Dar es Salaam, Tanzania; National Institute for Medical Research, Dar es Salaam, Tanzania; Harvard T.H Chan School of Public Health, Boston, MA, USA; Faculty of Pharmaceutical Sciences, Monash University, Melbourne, Australia; Department of Biochemistry, Kampala International University in Tanzania, Dar es Salaam, Tanzania

**Keywords:** Prevalence of malaria infections, risks of malaria infections, micro-geographic level, *Plasmodium falciparum*, Tanzania

## Abstract

**Background:** Despite a recent reduction in malaria morbidity and mortality, the disease remains a major cause of morbidity and mortality in Tanzania. However, the malaria burden is heterogeneous with a higher burden in some regions compared to others, suggesting that stratification of malaria burden and risk/predictors of infections is critical to guide the proper use of the current and future interventions. This study assessed the prevalence and predictors of /risk factors associated with malaria infections at micro-geographic levels in three villages of Muheza district, Tanga region, north-eastern Tanzania.

**Methods:** A cross-sectional community survey was conducted in three villages; Magoda, Mpapayu, and Mamboleo in Muheza district, Tanga region, north-eastern Tanzania in June 2021. Participants’ demographic, anthropometric, clinical, and malaria protection data were collected during the survey and combined with census data collected in 2013 including housing conditions and socio-economic status (SES). Finger prick blood samples were taken for parasite detection using both microscopy and rapid diagnostic tests (RDT). A generalised estimating equation (GEE) was used to determine the association between the prevalence and predictors/risk factors of malaria infections.

**Results:** The survey covered 1,134 individuals from 380 households and most of them (95.2%) reported that they slept under bed nets the night before the survey. By both microscopy and RDT, the prevalence of malaria infections was 19.2% and 24.3%, respectively. The prevalence was significantly higher among school children (aged >5 – 15 years, with 27.3% by microscopy and 37.6% by RDTs) compared to under-fives and adults (aged ≥15 years (p<0.001)). Individuals with a history of fever within 48 hours before the survey and those with fever at presentation (auxiliary temperature ≥37.5^0^C) were more likely to have malaria infections by microscopy (AOR = 1.16; 95% CI, 1.10 – 1.22; p<0.001) and RDTs (AOR = 1.18; 95% CI, 1.13 – 1.23; p<0.001). Participants with high SES and living in good houses (with closed eaves and/or closed windows) were less likely to be infected by malaria parasites as detected by microscopy (AOR =0.97; 95% CI, 0.92 - 1.02; p=0.205) and RDTs (AOR = 0.91; 95% CI, 0.85 - 0.97; p<0.001). Among the three villages, the prevalence of malaria by microscopy ranged from 14.7% to 24.6% and varied significantly but without any clear patterns across villages indicating high heterogeneity and random distribution of malaria at micro-geographic levels (p=0.001).

**Conclusion:** The villages had high prevalence and predictor/risk factors risk of malaria infections including age, sex (male), fever, SES, and housing conditions. High prevalence and risk were among school children (aged ≥5 - 14 years), males, individuals with low SES and a history of fever within 48 hours before the survey, or fever at presentation (with auxiliary temperature ≥37.5^0^C). The prevalence varied over short distances at micro-geographic levels suggesting that causes of such variations need to be established and considered when designing and implementing targeted malaria control interventions.

## BACKGROUND

Due to the scale-up and intensified control interventions in the past two decades, Tanzania and other Sub-Saharan African (SSA) countries succeeded in significantly reducing malaria morbidity and mortality between 2000 and 2019 [1]. The main interventions that have been used for malaria control by malaria endemic countries (and Tanzania as well) targeted mainly vectors, parasites (case management), and others. For vector control, insecticide-treated bed nets (ITNs) or long-lasting insecticidal nets (LLINs), indoor residual spraying (IRS), and larva source management (LSM) have been the core methods while case management interventions include parasitological diagnosis using rapid diagnostic tests (RDTs) and prompt treatment with artemisinin-based combination therapies (ACTs). Other interventions included chemoprevention such as intermittent preventive treatment (IPTp) in pregnant women [2], together with other preventive treatments as recommended by the World Health Organization (WHO) [3]. However, global malaria control and elimination efforts currently face major challenges such as insecticide [4] and antimalarial resistance [5], emergence and spread of parasites with deletions of histidine-rich protein 2/3 (*hrp2/3*) gene which cannot be detected by the HRP2 - mRDTs [6] and invasive vector species, Anopheles stephensi [7].

In Tanzania, malaria cases have declined tremendously over the past two decades from >18 million per year in the 2000s to less than 7 million in 2022, and deaths decreased from about 100,000 to less than 7,000 over the same period [8]. Despite these remarkable achievements, malaria is still a major public health threat and Tanzania was among the four countries (together with Nigeria, the Democratic Republic of Congo, and Niger) from SSA which contributed over half of all global malaria deaths in 2023 [9]. In addition, the malaria burden in Tanzania has become heterogeneous with some regions recording a high burden compared to other areas [10]. For instance, malaria prevalence within the country has declined from 18.1% in 2007/2008 to 7.9% in 2022 [11,12] with a remarkable epidemiological transition from meso-endemic to hypo-endemic levels in most parts of the country, resulting in distinct variations across and within regions and/or councils [13]. Some parts of Mainland Tanzania especially the north-western and southern regions have reported persistently high prevalence (some with ≥50%) among school children (aged ≥5 to 15 years) while the rest of the country has maintained low/very low prevalence over the past 20 years [14–16]. These epidemiological changes have been attributed to multiple factors including scaled-up interventions such as vector control with ITN/LLIN and IRS, and improved case management involving testing using RDTs followed by treatment of positive cases using ACTs. Others included chemopreventive [17,18] and non-intervention factors such as climate, physical environment, and socio-economic development [19,20].

Malaria infections in different geographical areas are characterised by variations in exposure to infective mosquitoes and risk factors associated with the infections [21–23], and these vary from one area to another at both micro and macro-geographic levels. For effective malaria control and the success of the ongoing elimination efforts, it is critical to establish with high precision the burden of malaria in each area targeted by the control efforts and identify the main predictors/risk factors associated with the local epidemiological pattern and transmission intensities of malaria. In Tanzania, the midterm review of the 2015 - 2020 National Malaria Control Programme’s (NMCP) national malaria strategic plan (NMSP) showed that the anticipated targets would not be met and thus strategic decisions and adjustments had to be made by the NMCP [24,25]. Together with its partners, NMCP revised its NMSP and initiated malaria stratification to establish the burden of the disease at the council levels to guide area-specific interventions and accelerate progress towards the country’s elimination target by 2030 [25]. In the 2020 malaria stratification, the country was divided into five epidemiological strata with councils and regions of high, moderate, low, and very low, and the urban stratum with 116/184 (63.1%) of the councils in high/moderate while the low/very low strata had 68/184 (36.9%) of all the councils [16]. The next round of stratification was done in 2022 and showed a slight increase of councils (70/184 (38.0%) located in low and very low malaria transmission [26]. Based on stratification, NMCP designed and has been implementing area-specific interventions with the goal of burden reduction in areas with high and moderate transmission and eliminating malaria in low/very transmission areas [25]. In addition, NMCP has promoted and intensified malaria surveillance, monitoring, and evaluation as a core intervention based on WHO recommendations [27].

While malaria burden stratification has been undertaken using both community parasite prevalence and routine health service data primarily over large geographic areas [28], some evidence indicates that spatial and temporal variations at micro-geographic levels also exist [29,30] but these have not been included in the initiatives pursued by NMCP in Tanzania. A recent study conducted in an area of high malaria transmission in north-western Tanzania highlighted marked differences in malaria prevalence within a small geographic area [31]. Other studies conducted within the country supported the same findings of high malaria prevalence with wide variations at micro-geographic levels [15,32]. High variation of malaria at micro-geographic levels could be due to the nature and distribution of risk factors such as breeding sites, housing conditions, elevation, and disparity in the availability, access, and utilisation of control interventions [33]. These and other risk factors may work singly or collectively to determine the prevalence of malaria in an area and if the factors are unevenly distributed within a particular geographic area, micro-geographic variations in the burden of malaria and intensities of transmission can be observed. Understanding the prevalence and risk factors associated with malaria infections at a micro-geographic level is crucial for designing effective control and prevention strategies. Micro-geographic studies provide a detailed analysis of malaria infections and their patterns and offer valuable insights into the specific factors contributing to the transmission of the disease within a limited geographical area. Therefore, this study assessed the prevalence and predictors of/risk factors associated with malaria infections at the micro-geographic level in three villages of Muheza district, north-eastern Tanzania.

## Methods

### Study area and design

This was a community-based cross-sectional survey (CSS) that was conducted in June 2021 and covered three villages of Magoda, Mamboleo, and Mpapayu in Muheza district, Tanga region, north-eastern Tanzania (Figure 1). It was conducted as part of the project on molecular surveillance of malaria in Tanzania (MSMT) which was established in 13 regions in 2021, before it was extended to cover all 26 regions of Mainland Tanzania in 2023 [34–36]. Muheza district is one of the nine districts of Tanga region in Tanzania which covers an area of 1,498 km^2^. The current and other villages of Muheza have been involved in extensive malaria research and other infectious diseases over the past four decades as described elsewhere [19,21,22]. According to the census update which was conducted in the study villages in May 2013 (and updated in September 2021), the three villages had a population of 2,934 individuals residing in 678 households (HHs) [22].

**Figure 1:**
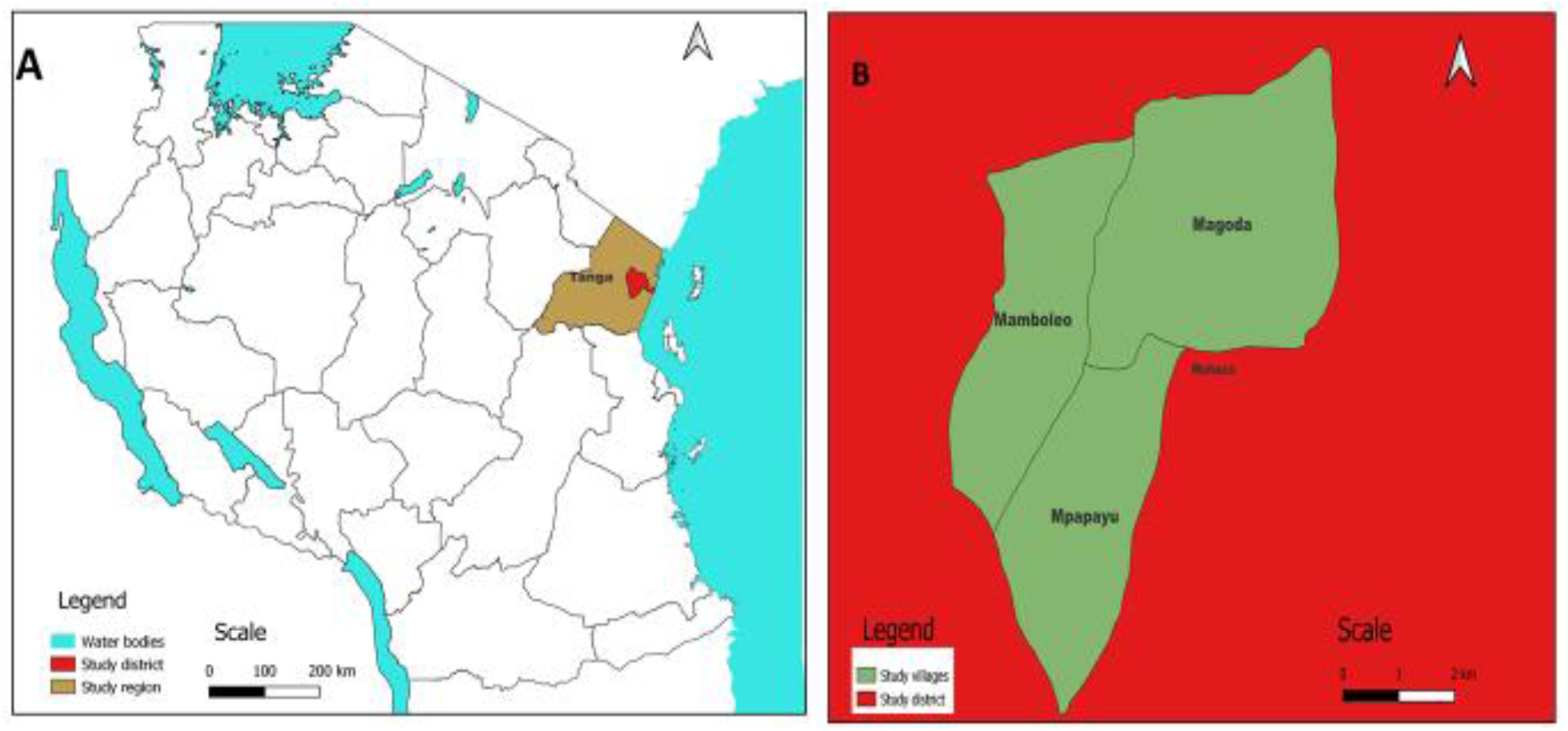
Maps of Tanzania showing the location of Tanga region (grey) and Muheza district (red) (A) and Study villages (right green) (B).

### Study population and recruitment of participants

This study targeted about 30% of all individuals aged >5 months from all HHs covered during the census done in 2013 (updated in 2021) using the MSMT protocol as described elsewhere [31]. All study participants were residents of the three villages in Muheza district (Magoda, Mamboleo, and Mpapayu) which are under the MSMT project. The studied individuals provided consent to participate in the study and signed an informed and/or accent form (for children 12 to 17 years old). With the support of village and hamlet leaders, information about the study was passed to each HH by a village sensitization team and all eligible individuals were invited to participate in the CSS. Each hamlet participated in the survey according to the schedule prepared by the data collection team. The team worked in each village for 2 - 3 days and any person who missed the visit in their respective village was advised to visit the team in the next village.

### Data collection procedures

There were two sources of data that were included in this study: household census data from the 2013 survey that was updated in December 2020 and CSS data. The household data from the household census [22] were collected by the research team by visiting each house to interview the head of the family and conduct observations of the different features of interest for the project. The data collected included household information, characteristics of the houses such as types and conditions of the houses (walls, roofing, floor and windows building materials status), the environment around the household (land use, vegetation cover, and breeding sites as well as large water bodies) and geo-referenced data of each household. The CSS data were collected during the CSS as described earlier and using a similar workflow with station-specific activities [31]. Eligibility criteria included age from 6 months and above, residence in the study village, and provision of informed consent. Individuals who were not residents of the study villages or did not meet the other criteria were not enrolled in the study but received medical care from the study clinical team. Consenting participants went through the registration process to confirm their identity and were assigned new numbers specific to the CSS. Thereafter, they were interviewed by trained researchers/research assistants to assess the use of any malaria control and preventive measures, and this was followed by anthropometric measurements (height, weight, and temperature). The next station was a mini-laboratory where finger prick was conducted by trained laboratory technicians or research nurses to obtain blood samples. Each participant was screened for malaria using RDTs and samples were collected for laboratory analysis including blood slides for detection, identification, and quantification of parasites by microscopy and dried blood spots (DBS) on filter papers. After sample collection, participants proceeded to the clinical section where thorough physical and clinical assessments were done by the study clinicians, to collect information on the history of fever, treatment history as well as treatment given before and during the survey. Participants who tested positive for malaria were treated according to the existing national guidelines [37] and all other conditions were also treated accordingly. Additionally, data including administrative boundary shape files were obtained from the National Bureau of Statistics [38] and used for mapping.

### Sample processing and generation of microscopy data

Samples collected as DBS were stored safely and later shipped to the laboratory for various analyses as reported elsewhere [36]. Blood smears were dried and thin smears were fixed using methanol. The smears were stained in the field with 5% Giemsa solution for 45 minutes, stored safely in slide folders and later transferred to the laboratory at the National Institute for Medical Research (NIMR) headquarters in Dar es Salaam for examination. Each sample was read under a high-power objective (100X magnification) by two independent microscopists blinded to each other’s results. In positive samples, asexual and sexual parasites were counted against 200 and 500 white blood cells (WBC), respectively. The parasite density was obtained by multiplying the number of parasites by 40 for asexual parasites and 16 sexual parasites (assuming each microliter of blood contained 8000 WBCs). Any smear was declared negative after examination of 200 high-power fields. For quality control, two qualified microscopists read all the slides independently, and parasite densities were calculated by averaging the two counts. Blood smears with discordant results (differences between the two microscopists in species detection, parasite density of >50% or the presence of parasites) were re-examined by a third, independent microscopist, and parasite density was calculated by averaging the two closest counts. Any other samples with discordant results were resolved by a team of three technicians who re-examined them at the same time as described elsewhere [39].

### Data management and analysis

The data were collected using a CSS questionnaire which was installed on tablets using Open Data Kit (ODK) software and stored on a central server located at the NIMR’s-Tanga Centre. All data were downloaded to a Microsoft Excel spreadsheet for quality check and cleaned for completeness against available data sources. Data analysis was performed using STATA version 17 (STATA Corp Inc., TX, USA). Descriptive statistics including frequencies and proportions were used to describe categorical variables while means and standard deviation or median with interquartile ranges were used to summarise continuous variables. The chi-square test was used to assess bivariate relationships between malaria prevalence and other categorical covariates. Generalised Estimating Equation (GEE) was used to assess risk factors associated with malaria infections [40]. Then, malaria prevalence was linked to geo-coordinates data to create maps using QGIS software. The distance from each participant’s HH to the nearest health facility, which is a dispensary located in Magoda village, and to the pond located near the three villages and any other potential breeding sites of mosquitoes was included in the analysis. The distance was categorised into low (0-1,000), moderate (1,000 - 2,000), and high (2,000 and above). This approach aimed to ascertain how such distance could influence the risk of infections at the micro-geographical level. The effects of different covariates in the delimited text layer file format were fitted into the QGIS map canvas to locate a breeding site and health facility within a map. Also, the prevalence of malaria in each village was added to the map canvas after each village’s malaria prevalence and fitted into the attribute table of a village map layer. Maps coordinates were generated by using World Geodetic System 1984 (WGS84) [41] and enhanced by the European Petroleum Survey Group (EPSG: 4326) as a database of coordinate system information and related documents on projections and datum [42]. The results showing the associations between independent and dependent variables were presented using odd ratios (OR) with 95% confidence intervals (CI). P-value ≤0.05 was considered statistically significant.

## Results

### Baseline characteristics of study participants

The 2021 CSS recruited 1,134 individuals with a mean age of 26.7 (SD=23.5) and residing in 380 HHs and all participants were assessed for malaria infections using RDTs and microscopy. The majority (53.3%) of participants were aged ≥15 years and most of them were females (57.1%), with males accounting for 42.9% of all participants. Over 75% of the participants were from HHs with open eaves and 47.8% had a history of fever within 48 hours before the survey while only 27 (2.4%) individuals had fever at presentation (axillary temperature ≥37.5^0^C). Most of the participants (95.2%) reported having slept under bed nets the night before the survey (Table 1).

**Table 1:**
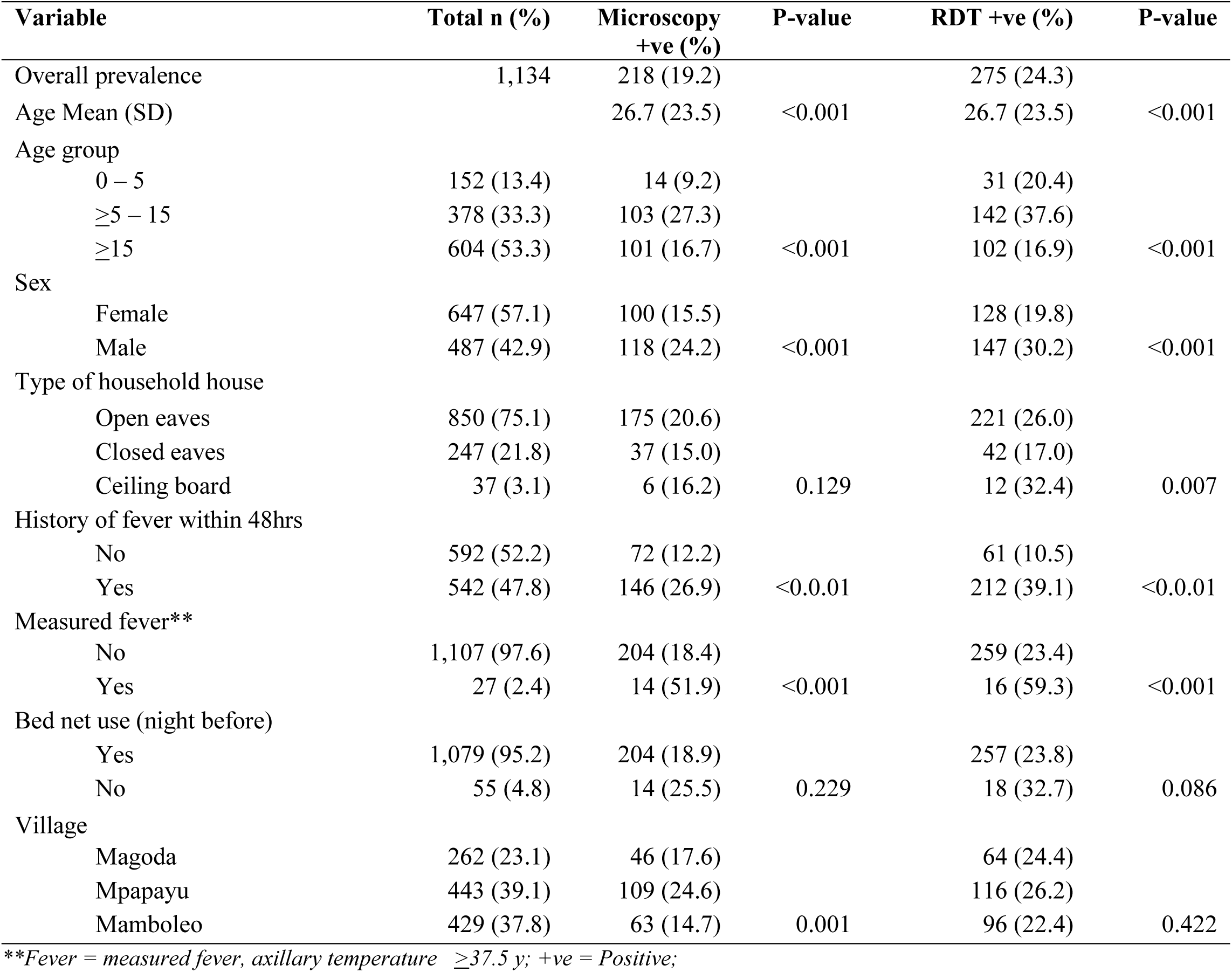
Baseline characteristics of study participants.

| Variable | Total n (%) | Microscopy<br>+ve (%) | P-value | RDT +ve (%) | P-value |
| --- | --- | --- | --- | --- | --- |
| Overall prevalence | 1,134 | 218 (19.2) |  | 275 (24.3) |  |
| Age Mean (SD) |  | 26.7 (23.5) | <0.001 | 26.7 (23.5) | <0.001 |
| Age group |  |  |  |  |  |
| 0 – 5 | 152 (13.4) | 14 (9.2) |  | 31 (20.4) |  |
| ≥5 – 15 | 378 (33.3) | 103 (27.3) |  | 142 (37.6) |  |
| ≥15 | 604 (53.3) | 101 (16.7) | <0.001 | 102 (16.9) | <0.001 |
| Sex |  |  |  |  |  |
| Female | 647 (57.1) | 100 (15.5) |  | 128 (19.8) |  |
| Male | 487 (42.9) | 118 (24.2) | <0.001 | 147 (30.2) | <0.001 |
| Type of household house |  |  |  |  |  |
| Open eaves | 850 (75.1) | 175 (20.6) |  | 221 (26.0) |  |
| Closed eaves | 247 (21.8) | 37 (15.0) |  | 42 (17.0) |  |
| Ceiling board | 37 (3.1) | 6 (16.2) | 0.129 | 12 (32.4) | 0.007 |
| History of fever within 48hrs |  |  |  |  |  |
| No | 592 (52.2) | 72 (12.2) |  | 61 (10.5) |  |
| Yes | 542 (47.8) | 146 (26.9) | <0.0.01 | 212 (39.1) | <0.0.01 |
| Measured fever** |  |  |  |  |  |
| No | 1,107 (97.6) | 204 (18.4) |  | 259 (23.4) |  |
| Yes | 27 (2.4) | 14 (51.9) | <0.001 | 16 (59.3) | <0.001 |
| Bed net use (night before) |  |  |  |  |  |
| Yes | 1,079 (95.2) | 204 (18.9) |  | 257 (23.8) |  |
| No | 55 (4.8) | 14 (25.5) | 0.229 | 18 (32.7) | 0.086 |
| Village |  |  |  |  |  |
| Magoda | 262 (23.1) | 46 (17.6) |  | 64 (24.4) |  |
| Mpapayu | 443 (39.1) | 109 (24.6) |  | 116 (26.2) |  |
| Mamboleo | 429 (37.8) | 63 (14.7) | 0.001 | 96 (22.4) | 0.422 |

\*\*Fever = measured fever, axillary temperature $\geq 37.5$ y; +ve = Positive;

### Malaria prevalence

The overall prevalence of malaria infections was 19.2% and 24.3% by microscopy and RDTs, respectively (Table 1). The prevalence was significantly higher among children aged >5 – 15 years (27.3% by microscopy and 37.6% by RDT) compared to under-fives (9.2% by microscopy and 20.4% by RDTs) and adults ≥15 years old (16.7% by microscopy and 16.9% by RDTs) (p-value<0.001). The prevalence was also significantly higher in males and among participants with a history of fever within 48 hours before the survey or with fever at presentation (axillary temperature ≥37.5^0^C) (p<0.001 for all comparisons). Individuals living in houses with open eaves had higher prevalence by both microscopy (20.6%) and RDTs (26.0%), but the differences were only significant for RDT results (Table 1).

### Risk factors of malaria infections in the study area

By both RDTs and microscopy, school children (aged ≥5 – 15 years) had a significantly higher risk of malaria infections before and after adjusting for potential covariates such as sex, type of house (presence or absence of eaves), history of fever within 48 hours and fever at presentation (RDTs; AOR = 1.18; 95% CI, 1.13 – 1.23; p<0.001; and microscopy AOR = 1.16; 95% CI, 1.10 – 1.22; p<0.001) (Table 2). Males were more likely to have malaria infections compared to female participants even after adjusting for age group, fever, and type of house (with open or closed eaves) (RDT results: AOR = 109; 95% CI: 1.03 - 1,14; p=0.001; and microscopy results: AOR = 1.06; 95% CI:1.02 – 1.11; p=0.005). Participants with a history of fever within 48 hours before the survey were also more likely to have the infections compared to those without fever (microscopy results: (AOR = 1.13; 95% CI: 1.08 – 1 .18; p<0.001; and RDT results: AOR = 1.28; 95% CI: 1.22 - 1.35; p<0.001). Similarly, participants with fever at presentation (axillary temperature ≥37.5^0^C) were more likely to have malaria infections compared to those without fever (microscopy results: AOR = 1.36; 95% CI: 1.13 – 1.63; p = 0.001; and RDT results: AOR = 1.35; 95% CI: 1.12 - 1.61; p=0.002). Study participants living in houses with closed eaves had a lower risk of malaria infection but the results were only significant for RDTs (microscopy results: AOR = 0.97; 95% CI:0.92 – 1.02; p=0.205; and RDT results; AOR = 0.91; 95%CI:0.85 – 0.97; p<0.001). The risk of malaria infections among individuals who did not sleep under bed-nets was similar to that of bed-net users (microscopy results: AOR = 1.05, 95% CI: 0.93 – 1.19, p=0.444 and RDT results: AOR = 1.02, 95%CI: 0.91 – 1.14, p=0.913), suggesting that non-users were potentially protected by bed-net users (Table 2).

**Table 2:** Risk of malaria infections in the three villages of Magoda, Mamboleo and Mpapayu.

| Variables | Microscopy |  |  |  | RDTs |  |  |  |
| --- | --- | --- | --- | --- | --- | --- | --- | --- |
|  | COR (95%CI) | P-value | AOR (95%CI) | P value | COR (95%CI) | P value | AOR (95%CI) | P value |
| Age group |  |  |  |  |  |  |  |  |
| 0 – 5 | 1 |  | 1 |  | 1 |  | 1 |  |
| ≥5 – 15 | 1.18(1.11–1.26) | <0.001 | 1.16(1.10–1.22) | <0.001 | 1.18(1.10–1.28) | <0.001 | 1.18(1.13–1.23) | <0.001 |
| ≥15 | 1.09(1.04–1.15) | 0.001 | 1.10(1.07–1.14) | <0.001 | 0.98(0.91–1.04) | 0.490 | 0.97(0.91–1.04) | 0.339 |
| Sex |  |  |  |  |  |  |  |  |
| Female | 1 |  | 1 |  | 1 |  | 1 |  |
| Male | 1.08(1.03–1.13) | 0.001 | 1.06(1.02–1.11) | 0.005 | 1.10(1.05–1.15) | <0.001 | 1.09(1.03–1.14) | 0.001 |
| History of fever within 48hrs |  |  |  |  |  |  |  |  |
| No | 1 |  | 1 |  | 1 |  | 1 |  |
| Yes | 1.15(1.10–1.20) | <0.001 | 1.13(1.08–1.18) | <0.001 | 1.31(1.25–1.38) | <0.001 | 1.28(1.22–1.35) | <0.001 |
| Measured fever** |  |  |  |  |  |  |  |  |
| No | 1 |  | 1 |  | 1 |  | 1 |  |
| Yes | 1.36(1.14–1.65) | 0.001 | 1.36(1.13–1.63) | 0.001 | 1.36(1.13–1.63) | 0.001 | 1.34(1.12–1.61) | 0.002 |
| Bed net use: |  |  |  |  |  |  |  |  |
| Slept under | 1 |  | 1 |  | 1 |  | 1 |  |
| Slept without | 1.09(0.96–1.23) | 0.189 | 1.02(0.91–1.14) | 0.913 | 1.08(0.95–1.22) | 0.248 | 1.05(0.93–1.19) | 0.444 |
| Type of household house |  |  |  |  |  |  |  |  |
| Open eaves | 1 |  | 1 |  | 1 |  | 1 |  |
| Closed eaves | 0.97(0.91–1.02) | 0.245 | 0.97(0.92–1.02) | 0.205 | 0.93(0.88–0.99) | 0.015 | 0.91(0.85–0.97) | <0.001 |
| Ceiling board | 0.98(0.85–1.12) | 0.775 | 0.72(0.86–1.11) | 0.723 | 1.10(0.93–1.30) | 0.277 | 0.99(0.90–1.08) | 0.880 |

*\*\*Measured fever/fever at presentation $\geq 37.5^{\circ}\text{C}$ on the day of survey; COR=Crude Odds Ratios; AOR=Adjusted Odds Ratios; CI=Confidence Interval*

### Micro-geographic differences in the prevalence of malaria infections in the study area

Despite their very close proximity (less than 1 km apart), the prevalence by microscopy varied significantly within the study villages (p=0.001) while the prevalence was not significant by RDT (p=0.422). The highest microscopy prevalence was observed in Mamboleo (24.60%) while by RDT, the highest was in Mpapayu (34.91). The lowest microscopy prevalence was in Mpapayu (14.69%) while RDT’s lowest prevalence (24.05) was reported in Magoda (Table 3). The spatial distribution of malaria infections within the three villages had no clear pattern suggesting that the risk of infections was randomly distributed (Figure 2).

**Figure 2:**
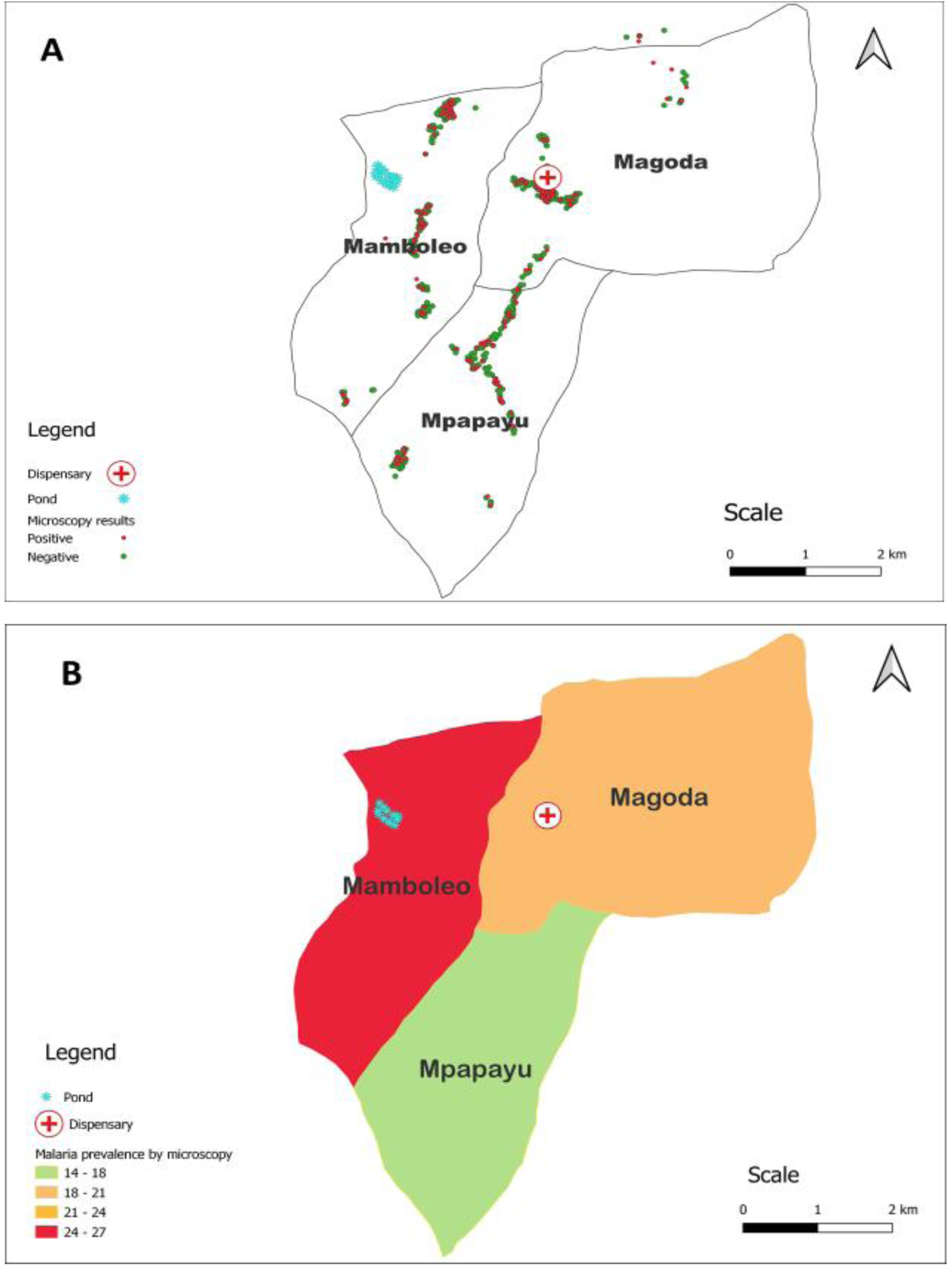
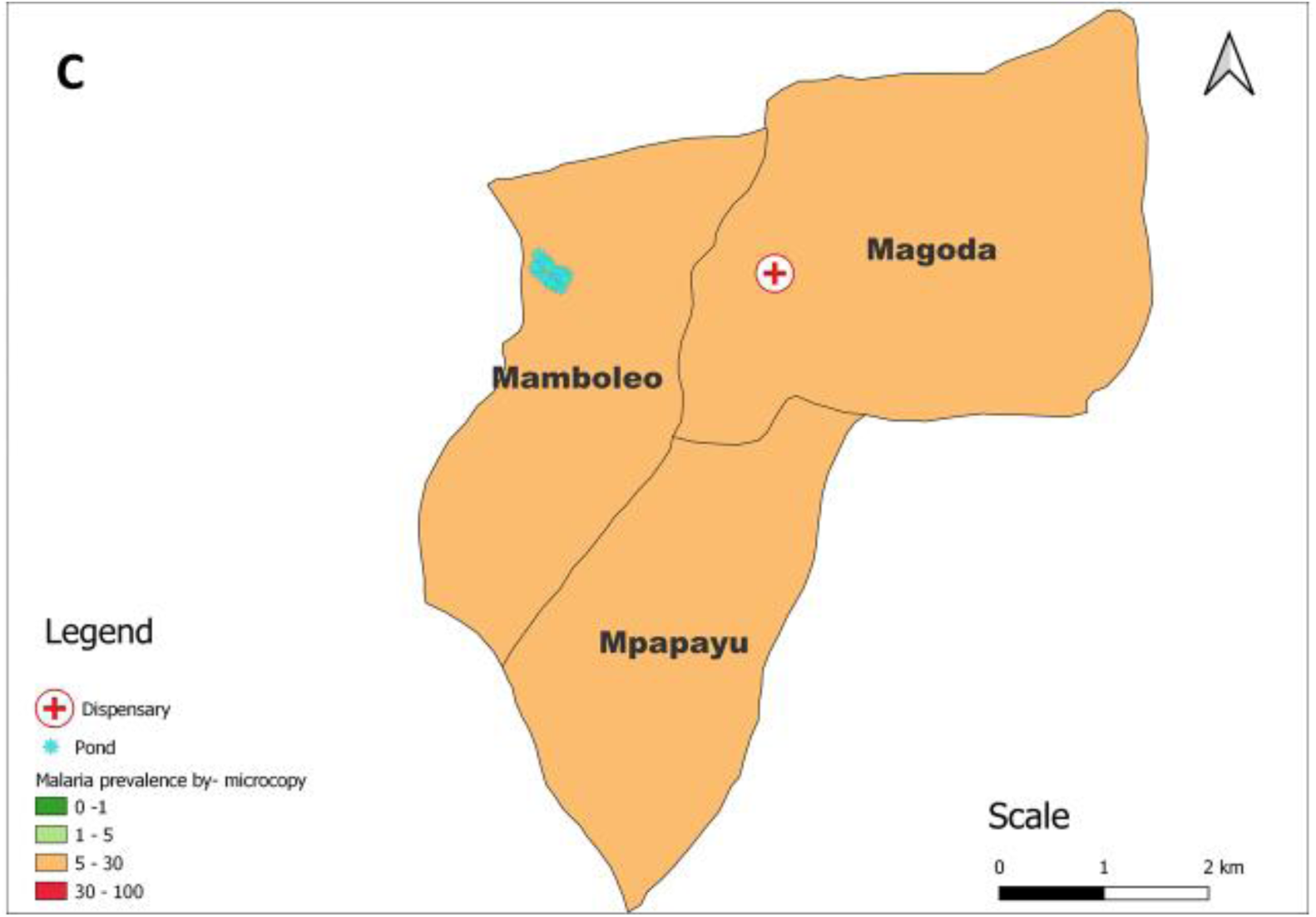
A map of the study villages showing the location of tested individuals and their results by microscopy (A), the average prevalence of malaria infections (B), and the prevalence of malaria infection stratification per NMCP strata (C).

**Table 3:** Prevalence of malaria infection among individuals by micro-geographical data.

| Variable | Total | Microscopy |  |  | RDTs |  |  |
| --- | --- | --- | --- | --- | --- | --- | --- |
|  |  | Yes (%) | No (%) | P-value | Yes (%) | No (%) | P-value |
| Dispensary |  |  |  |  |  |  |  |
| Low (0–1,000) | 324 (28.57) | 65 (20.06) | 259 (79.94) |  | 84 (25.92) | 240 (74.07) |  |
| Moderate (1,000–2,000) | 347 (30.60) | 60 (17.29) | 287 (82.71) |  | 81 (23.34) | 266 (76.66) |  |
| High ($\geq 2,000$) | 463 (40.83) | 93 (20.09) | 370 (79.91) | 0.548 | 110 (23.76) | 353 (76.24) | 0.701 |
| Pond |  |  |  |  |  |  |  |
| Low (0–1,000) | 155 (13.67) | 26 (16.77) | 129 (83.23) |  | 32 (20.65) | 123 (79.35) |  |
| Moderate (1,000–2,000) | 267 (23.54) | 46 (17.23) | 221 (82.77) |  | 67 (25.09) | 200 (74.91) |  |
| High ($\geq 2,000$) | 712 (62.79) | 146 (20.51) | 566 (79.49) | 0.361 | 176 (24.72) | 536 (75.25) | 0.526 |
| Village |  |  |  |  |  |  |  |
| Magoda | 262 (23.10) | 46 (17.56) | 216 (82.44) |  | 63 (24.05) | 199 (75.95) |  |
| Mamboleo | 443 (39.07) | 109 (24.60) | 334 (75.40) |  | 116 (26.19) | 327 (73.81) |  |
| Mpapayu | 429 (37.83) | 63 (14.69) | 366 (85.31) | 0.001 | 96 (34.91) | 333 (77.62) | 0.422 |

This study also explored the prevalence and the likelihood of malaria infection by assessing some of the micro-geographic features around and within the studied villages. The features include distance from the health facility (Magoda dispensary) and distance from the large pond which is located in Mamboleo village. Both crude and adjusted analyses showed the same results as the likelihood of malaria infections was not significantly influenced by the presence of these micro-geographic features. The likelihood of malaria infection by microscopy was significantly higher in Mamboleo village where the pond is located (AOR = 1.63; 95%CI:1.10 - 2.41; p=0.015) than in the other two villages (Tables 4).

**Table 4:** Univariate and multivariate association of malaria infections by micro-geographical factors.

| Distance (m) | Microscopy |  |  |  | RDTs |  |  |  |
| --- | --- | --- | --- | --- | --- | --- | --- | --- |
|  | COR (95%CI) | P-value | AOR (95%CI) | P value | COR (95%CI) | P value | AOR (95%CI) | P value |
| Dispensary |  |  |  |  |  |  |  |  |
| Low (0–1,000) | 1 |  | 1 |  | 1 |  | 1 |  |
| Moderate (1,000–2,000) | 0.83(0.56–1.23) | 0.357 | 0.97(0.59–1.58) | 0.891 | 0.87(0.61–1.24) | 0.438 | 0.91(0.58–1.42) | 0.670 |
| High (≥2,000) | 1.00(0.70–1.43) | 0.993 | 1.03(0.72–1.48) | 0.856 | 0.89(0.64–1.24) | 0.488 | 0.90(0.64–1.26) | 0.534 |
| Pond |  |  |  |  |  |  |  |  |
| Low (0–1,000) | 1 |  | 1 |  | 1 |  | 1 |  |
| Moderate (1,000–2,000) | 1.03(0.61–1.75) | 0.905 | 1.03(0.60–1.75) | 0.923 | 1.29(0.80–2.08) | 0.299 | 1.28(0.79–2.06) | 0.320 |
| High (≥2,000) | 1.28(0.81–2.03) | 0.292 | 1.24(0.72–2.14) | 0.437 | 1.26(0.83–1.93) | 0.283 | 1.21(0.73–2.01) | 0.449 |
| Village |  |  |  |  |  |  |  |  |
| Magoda | 1 |  | 1 |  | 1 |  | 1 |  |
| Mamboleo | 1.53(1.04–2.25) | 0.030 | 1.63(1.10–2.41) | 0.015 | 1.12(0.79–1.60) | 0.528 | 1.15(0.80–1.65) | 0.446 |
| Mpapayu | 0.81(0.53 – 1.22) | 0.316 | 0.81(0.53–1.24) | 0.331 | 0.92(0.63–1.31) | 0.613 | 0.93(0.64–1.34) | 0.689 |

## Discussion

This study was conducted to determine the prevalence and predictors of/risk factors associated with micro-geographic variations of malaria infections among individuals living in areas with declining malaria transmission in the Muheza district. The study villages and other parts of the district have experienced a significant decline in malaria burden from a prevalence of over 80.0% in individuals of all age groups in the 1990s to about 13.0% in 2017 [19]. Previous studies also showed that the burden of malaria shifted from under-fives to school children and was significantly influenced by the amount of rainfall together with other factors that could not be established [19]. This study builds on previous studies done in these and other villages of Muheza district and aims to provide more evidence to support NMCP’s efforts of undertaking micro-stratification of malaria and targeting interventions to persistent foci and vulnerable groups. The findings of this study indicate that the prevalence and risk of malaria were higher in school children (aged between ≥5 and 15 years), males, participants with fever within 48 hours before the survey, and those with fever at presentation (with axillary temperature ≥37.5^0^C), low SES and living in poorly constructed houses. Thus, these and other predictors/risk factors of malaria infections, which are yet to be identified need to be properly collated and targeted by the NMCP in the ongoing efforts to eliminate malaria by 2030.

In the current study, the overall prevalence was 19.2% by microscopy and 24.3% by RDTs which was higher than what was reported in previous surveys where the lowest prevalence was ≤7.2% in 2012, but increased to over 31.0% in 2015 and declined to 13.0% in 2017 [19], suggesting an unstable transmission patterns in these villages. In the last two decades, most endemic countries including those in the WHO-Afro region scaled-up malaria interventions and started to experience a decline in the disease burden specifically among under-fives but with a remarkable shift resulting in a higher burden of malaria in school children. These major epidemiological transitions have been attributed to scaled-up interventions such as ITNs/LLINs and improved case management based on prompt diagnosis and treatment of infections with effective antimalarial drugs [43]. Other interventions such as chemoprevention in pregnant women (IPTp and infants (IPTi), and seasonal malaria chemoprevention (SMC) have also been linked to these changes [44]. However, studies have shown that the changes in malaria epidemiology including variability of transmission intensities can be influenced by both biotic and abiotic factors such as physical environment, climate, vegetation, and human activities [20,45]. A study that was conducted in the villages covered in this study showed that the prevalence of malaria was significantly affected by the amount of rainfall with high prevalence in years that had excess rains above the average rainfall of the 30 years. That study also showed that other factors yet to be determined could have potentially played an important role in the trends of malaria over the past four decades with the unstable transmission patterns, shown by the increase which was followed by a decrease of prevalence from 2012 to 2017 [19]. Studies conducted elsewhere have also shown that the burden and transmission of malaria largely depend on different factors such as low SES (poverty), poor housing conditions, human activities that create more breeding sites (such as irrigation schemes or construction projects), h, and access to and utilisation of malaria interventions that need to be considered in designing and implementing malaria control/elimination strategies [46–49].

The decline in the malaria burden in under-fives and an increase in school children has been consistently reported and documented in the past two decades, and this is significantly attributed to recent interventions for malaria control. Studies conducted in different countries reported a decline in the prevalence of malaria infections in under-fives while the prevalence in school children either increased or had a slight decline between 2000 and 2023 [50,51]. These findings showing a high prevalence of malaria in school children are consistent with those of other studies that were conducted in Tanzania and Uganda and reported a high prevalence of malaria in this group[52–54]. This provides evidence to NMCP to support targeting school children with specific interventions aimed at shrinking the transmission reservoir in this group where most infections are asymptomatic, as the country makes progress towards malaria elimination by 2030.

Based on the findings of high malaria burden in school children and a decline of malaria prevalence in under-fives, different countries are targeting these groups differently with malaria interventions. For instance, in Tanzania, school children’s interventions include biannual parasitological surveys and free bed nets through the school net programme [55], while a plan is underway to launch intermittent preventive treatment (IPTsc) [52] to reduce the malaria burden in this group and prevent them from sustaining and perpetuating transmission. For under-fives, the government still implements the scheme of providing bed nets through the ANC clinics and also free health care that covers case management services, focusing on prompt diagnosis using RDTs and effective treatment for malaria using ACTs [56]. Despite these initiatives, the burden of malaria is still higher in school children compared to other groups [31] and the reasons for this are not known and urgent measures are required to generate more evidence and factors responsible for this pattern. In addition, efforts and strategies are needed to fully assess, understand, and support NMCP to implement appropriate interventions to reduce the burden of malaria in this group.

This study also showed that the prevalence and risk of malaria infections were higher among males compared to females. In this study, additional review of the data showed that the highest prevalence among males was observed in boys aged 5 - 15 years followed by those aged >15yrs. Similarly, the studies conducted elsewhere including in Cameroon, Nigeria, and Uganda also showed that the prevalence of malaria infections was higher among males [57–59]. High prevalence in males may be attributed to several factors including the habit of staying outdoors very late at night before going to bed covered with bed nets, especially for the adult males. It has also been linked to drinking behaviour whereby men take alcohol in open areas at night and also other socio-economic activities during the night which increase exposure to potentially infectious mosquito bites [60]. Studies have also shown that females tend to be more aware of various methods used for malaria prevention and control and use them more often than males, with higher knowledge and use of interventions such as ITNs and SP for IPTp [61,62]. Females also have more awareness of and high health-seeking behaviour compared to men due to their roles as caretakers of children and their families [63]. Although these factors may have contributed to the potential lower risk of malaria infections among females, some studies have shown contradictory findings with reports of the high prevalence of malaria in females, as recently shown in one of the villages of Kyerwa district in Kagera region [31] and another from Nyasa district in Ruvuma region [64]. Thus, more studies may be needed to confirm these findings and provide evidence of the high vulnerability of males to malaria infections so that this group can also be targeted with malaria interventions such as those aimed at reducing the overall vector abundance and/or protecting them from outdoor biting

The results from this study revealed that prevalence of malaria infections was twice as much among individuals who had a fever at presentation or had a history of fever within 48 hours before the survey compared to those without a fever. This is consistent with studies conducted elsewhere that reported high rates of infections in febrile individuals [65]. Fever is therefore considered a strong predictor of malaria infection particularly in areas of high transmission intensities [66]. One of the primary ways to determine the effectiveness of antipyretic therapy is to investigate the impact of temperature on asexual phase parasite growth and reproduction [67]. In Tanzania, the NMCP case management policy emphasizes the parasitological confirmation of malaria before treatment in patients of all ages [25]. However, some febrile individuals fail to seek healthcare services potentially due to accessibility and/or affordability problems such as long distances to health facilities, transport costs, and healthcare service costs, as reported elsewhere [46,68]. Therefore, the study proposes that the community firstly should seek malaria confirmation through testing before taking any malaria medications.

Housing conditions were shown to be an important predictor of malaria infection in this study. It was shown that individuals who were living in better houses with closed eaves, and with high SES had a lower risk of malaria infections. This is consistent with findings from other studies that recommended improvement of housing conditions [69,70] and reducing poverty in the fight against malaria [71]. Therefore, efforts to fight against malaria should also include increased poverty reduction campaigns and education for communities to enable them to build better houses with closed eaves, corrugated iron sheets, and cement walls [72]. However, adopting this strategy to reduce and control malaria mainly depends on the government’s and global partners’ strategies to enhance economic growth and ultimately improve the SES of the families. Previous studies showed that apart from their inability to afford better or improved houses, poor families also fail to adopt and effectively use other malaria interventions (such as access to better health care services, and ownership and use of bed nets) leaving them at increased risk of the disease as well as perpetuating the transmission in the community [73–75]. Therefore, specific interventions are urgently needed to target poor families in the fight against malaria. Such interventions should include increased access to different interventions such as bed nets, health care services for prompt and effective case management, chemoprevention, and behaviour change communication (BCC).

In the study areas, most of the participants (over 96%) reported owning and using bed nets and the prevalence and risk of malaria infections were similar in those with as well as without/not using bed nets. Studies have shown that with high bed net coverage, even non-users and the entire community may be protected against malaria because high coverage of ITN/LLINs in the community and their use lead to a reduction of the number of mosquitoes, their length of survival and eventually their ability to transmit malaria [76,77]. However, given the recent campaigns that aimed to attain universal coverage and use of bed nets in Tanzania, the reasons for these few individuals not owning and using bed nets are not clearly known and will need to be explored in future studies. It should also be noted that this data was obtained through self-reporting by participants and could therefore suffer from information and recall bias [78]. However, some studies have shown that households with poor economic status was a major reason for not owning and using bed nets as a protection against mosquito bites and malaria infection [79,80], though this is not observed in our study. Even if this relation is true either or not, there is still a need to develop ITN distribution mechanisms and training activities that will consider socio-economic disparities in bed net ownership. Such initiatives are critical and should target vulnerable groups to enhance the progress toward the reduction of the malaria burden in the entire community and reach the elimination targets.

The study also looked at some important features that could be associated with the variation of prevalence and predictors/risk of malaria infections at micro-geographic levels in these villages located next to each other (Figure 2). In each village, the prevalence was not significantly different in the sub-villages (the lowest administrative structure in Tanzania) suggesting that the risk factors could be similar at the village level. However, the prevalence and predictors/risk of malaria infections were unevenly distributed in the three villages. This indicates the presence of heterogeneity among villages and the importance of micro-geographic studies to provide more information about malaria burden at the granular level and possible causes of these patterns. In the current study, the prevalence of malaria infections ranged from 15% to 25% by microscopy and 24% and 35% for RDTs, and these variations were not significantly associated with the key features studied, the location of the dispensary, and a big pond. These findings are in contrast to what was reported by NMCP in 2020 and others in 2022 that Muheza district council including the study villages had an overall high prevalence which was 30% and above [25,26,28]. Among the study villages, Mamboleo had a slightly higher prevalence of malaria infections than the other two villages, possibly due to the presence of the pond within the village. However, the relationship between pond availability with malaria infection was not statistically significant. These findings suggest that more studies are required to tease out the different features that might be associated with persistent and unstable transmission patterns in these villages which have been studied since 1992.

The study was conducted in only three villages of Muheza district, Tanzania, which may not be a good representation of other areas.

## Conclusion

There was high prevalence in the study villages despite a decline reported up to 2012 and both the prevalence and predictors/risk of malaria infections were higher in schoolchildren, males, and individuals with a history of fever within 48 hours before the study, and those with fever at presentation (with axillary temperature ≥37.5^0^C). It was also revealed that malaria prevalence varied over a short distance at micro-geographic levels as the prevalence was significantly different among the three villages, despite their close proximity. This suggests that the causes of these variations need to be identified and addressed to reach the goal of malaria elimination by 2030. Moreover, these findings showed high coverage of bed nets (over 90 percent) but still the prevalence of malaria was high in these villages compared to previous studies which reported a decline and low prevalence. Thus, more studies including social anthropological surveys are urgently needed to reveal the reasons behind the persistence of malaria transmission in the study area, despite high coverage of interventions such as bed nets and effective case management.

## Data Availability

The dataset of this study is available from the corresponding author upon a reasonable request from the corresponding author together with an institutional approval by NIMR and signing of a data transfer agreement (DTA) between the donor and recipient.

## Declarations

### Ethical clearance

The ethical approval for conducting this study was obtained from the Medical Research Coordinating Committee (MRCC) of the National Institute for Medical Research (NIMR) in Tanzania. Permission to conduct the study in the villages was provided by the President’s Office, Regional Administration and Local Government Authority (PO-RALG), and health authorities of Tanga regional, Muheza district, and village authorities. Informed consent/assent was sought and obtained before conducting the study, from each participant or parents/legal guardians for children. Permission to publish this paper was sought and given by the Director General of the NIMR.

### Consent for publication

Not applicable.

### Competing interests

All authors declare that they have no competing interests.

### Funding

This work was supported in full by the Bill & Melinda Gates Foundation [grant number 002202]. Under the grant conditions of the Foundation, a Creative Commons Attribution 4.0 Generic License has already been assigned to the Author Accepted Manuscript version that might arise from this submission.

### Author’s contributions

DPC, ESKand DSI designed and conceptualised the study and revised the manuscript. All authors took part in data collection under the supervision of DSI, ESK, and VWM. DPC and FF managed and analysed data. FF, MDS, CIM, JBT, and RAM critically revised the manuscript. DPC developed the manuscript with the support of DSI and VWM. All authors read and approved the final version of the manuscript.

## Acknowledgment

The authors convey warm and cordial thanks to all study participants for their readiness to participate and take part in this study. They highly appreciate the support of village leaders, and district and regional medical authorities which was rendered to the study team during the entire study period. The study was executed by a committed study team including Gineson Nkya, Neema Barua, Francis Chambo, August Nyaki, Juma Akida, Athanas Mhina, Salome Simba, Dativa Pereus, Tilaus Gustav, Hatibu Athuman, Gasper Lugela, Richard Makono, Ildephonce Mathias, Arison Ekoni, Twalipo Mponzi, Denis Byakuzana, Pendael Nasary, Thomas Semdoe and Halfan Mwanga. The devoted support of NIMR management and staff is highly acknowledged. The entire study team and authors received technical and logistics support from partners at Brown University, the University of North Carolina at Chapel Hill, the CDC Foundation, and the Bill and Melinda Gates Foundation team. This support is highly appreciated.

## Availability of data and materials

The dataset of this study is available from the corresponding author upon a reasonable request from the corresponding author together with institutional approval by NIMR and the signing of a data transfer agreement (DTA) between the donor and recipient.

## Abbreviations and acronyms

ACT: Artemisinin-based combination therapy
DHIS2: District Health Information Systems 2
GEE: Generalised Estimating Equations
IPTi: Intermittent preventive treatment in infants
IPTp: Intermittent preventive treatment in pregnant women
IRS: Indoor residual spraying
ITNs: Insecticide-treated bed nets
LSM: Larval source management
MoH: Ministry of Health
MRCC: Medical Research Coordinating Committee
RDT: malaria rapid diagnostic test
MSMT: Molecular Surveillance of Malaria in Tanzania
NBS: National Bureau of Statistics
NMCP: National Malaria Control Program
ODK: Open Data Kit software
PCA: Principal component analysis
PO-RALG: President’s Office, Regional Administration and Local Government Authority
QGIS: Quantum Geographical Information System
SSA: Sub-Saharan Africa
TMIS: Tanzania Malaria Indicator Surveys
WHO: World Health Organisation

## Notes

### Competing Interest Statement

The authors have declared no competing interest.

### Author Declarations

The ethical approval for conducting this study was obtained from the Medical Research Coordinating Committee of the National Institute for Medical Research in Tanzania.

